# Development of risk prediction models for preterm delivery in a rural setting in Ethiopia

**DOI:** 10.1101/2022.11.04.22281948

**Authors:** Clara Pons-Duran, Bryan Wilder, Bezawit Mesfin Hunegnaw, Sebastien Haneuse, Frederick G. B. Goddard, Delayehu Bekele, Grace J. Chan

## Abstract

**Background:** Preterm birth complications are the leading causes of death among children under five years. A key practical challenge, however, is the inability to accurately identify pregnancies that are at high risk of preterm delivery, especially in resource-limited settings where there is limited availability of biomarkers assessment.

**Methods:** We evaluated whether risk of preterm delivery can be predicted using available data from a pregnancy and birth cohort in Amhara region, Ethiopia. All participants were enrolled in the cohort between December 2018 and March 2020. The study outcome was preterm delivery, defined as any delivery occurring before week 37 of gestation regardless of vital status of the fetus or neonate. A range of sociodemographic, clinical, environmental, and pregnancy-related factors were considered as potential inputs. Cox and accelerated failure time models, and decision tree ensembles were used to predict risk of preterm delivery. Model discrimination was estimated using the area-under-the-curve (AUC). Additionally, the conditional distributions of cervical length (CL) and fetal fibronectin (FFN) were simulated to ascertain whether those factors could improve model performance.

**Results:** A total of 2493 pregnancies were included. Of those, 138 women were censored due to loss-to-follow-up before delivery. Overall, predictive performance of models was poor. The AUC was highest for the tree ensemble classifier (0.60, 95%CI [0.57, 0.63]). When models were calibrated so that 90% of women who experienced a preterm delivery were classified as high risk, at least 75% of those classified as high risk did not experience the outcome. The simulation of CL and FFN distributions did not significantly improve models’ performance.

**Conclusions:** Prediction of preterm delivery remains a major challenge. In resource-limited settings, predicting high-risk deliveries would not only save lives, but also inform resource allocation. It may not be possible to accurately predict risk of preterm delivery without investing in novel technologies to identify genetic factors, immunological biomarkers or the expression of specific proteins.

## INTRODUCTION

Globally, almost 15 million babies are born preterm before 37 weeks of gestation each year [1]. A variety of factors are known to be associated with risk of preterm birth, including obstetrics history, anthropometric measurements, infections, ultrasound measurements and biological and genetic markers [2,3]. Accurate prediction tools to identify women at an increased risk of preterm delivery would allow policy makers, practitioners and researchers to target interventions designed to reduce preterm deliveries. Some studies conducted in high income settings developed predictive models to classify women based on their risk for preterm birth considering multiple maternal characteristics [4,5]. Their discriminative performance was modest (area under the receiving operator characteristic curve (AUC) from 0.62 to 0.70), with generally lower performance when performing external validation [5].

Targeting interventions to high-risk pregnancies is a critical challenge because of the lack of accurate prediction tools. Some published models were developed for women with *a priori* known risk factors like preterm labor or multiple pregnancy [6-8]. Other models used predictors that are not readily available in resource-limited settings such as cervical length (CL), bacterial vaginosis, fetal fibronectin (FFN), cytokine concentration and other biomarkers [9-11].

Additionally, to our knowledge, no published model handled competing risks of stillbirth, or considered a combined outcome of preterm delivery regardless of vital status of the fetus or neonate. However, preterm and stillbirth share common causes and risk factors, and it is likely that the biological mechanisms that trigger preterm labor or rupture of membranes may lead to the delivery of a preterm stillborn in extreme cases [12,13]. Overall, there is a gap in the development of prediction tools that are accurate and applicable to the general population, with and without *a priori* risk, especially in low-resource countries where data on biomarkers that could contribute to improve model performance are not commonly available.

Most preterm birth cases occur among women without known risk factors [14,15]. Limited availability of promising biomarkers to predict preterm birth in low-resource settings make it critical to develop context-specific predictive tools. We use large datasets from the Birhan pregnancy cohort [16], to test whether it is possible to predict risk of preterm delivery in rural Ethiopia. Additionally, we aim to ascertain whether it would be effective to invest in the collection of known key predictors such as CL or FFN to improve accuracy of predictions by simulating the conditional distribution of those two factors.

## METHODS

### Study design and setting

We conducted a cohort study in the Birhan field site, including 16 villages in Amhara region, Ethiopia, covering a population of 77,766, to estimate morbidity and mortality outcomes among 17,108 women of reproductive age and 8,554 children under-five with house-to-house surveillance every three months. The site is a platform for community and facility-based research and training that was established in 2018, with a focus on maternal and child health [17]. Nested in the site is an open pregnancy and birth cohort that enrolls approximately 2,000 pregnant women and their newborns per year with rigorous longitudinal follow-up over the first two years of life and household data linked with health facility information [16]. The catchment area is rural and semi-urban, covers both highland and lowland areas, and includes two different districts, Angolela Tera, and Kewet/Shewa Robit.

We used data from the Birhan Health and Demographic Surveillance System (HDSS) and the nested pregnancy and birth cohort, Birhan Maternal and Child Health (MCH) cohort to develop a series of risk prediction models for preterm delivery [16,17]. The HDSS provides estimates and trends of health and demographic outcomes including morbidity among women of reproductive age and children under two years and births, deaths, marriages, and migration in the entire population [17]. The pregnancy and birth cohort, generates evidence on pregnancy, birth, and child outcomes using clinical and epidemiological data at both the community and health facility level [16].

### Study participants

The sample for this study included women enrolled during pregnancy in the MCH cohort between December 2018 and March 2020. They were followed-up in home and facility visits through delivery. Women who were enrolled during pregnancy and were followed up beyond 28 weeks of gestation were included in the study. This gestational age cut-off was used because in Ethiopia stillbirths are considered ≥28 weeks. We excluded newborns with implausible gestational ages at birth: <28 weeks due to the definition of stillbirth, and ≥46 weeks.

### Study variables and definitions

The outcome of this study was *preterm delivery*, a composite indicator defined as any delivery occurring before 37 completed weeks of gestation, regardless of vital status of the fetus or neonate. This includes both *preterm births* (live birth prior to completion of week 37 of gestation) [18], as well as *stillbirths* (any fetal death after 28 completed weeks of gestation) [19] which occurred before 37 weeks of gestation.

Gestational age was estimated using the best available method from ultrasound measurements, reported date of last menstrual period, fundal height or maternal recall of gestational age in months. Detailed information on these estimations can be found elsewhere [20].

The selection of potential predictors was guided by literature review and expert knowledge from study obstetricians and pediatricians. Predictors with low prevalence rates in the sample (rare events with ≤ 5 cases) were dropped. Over 70 socio-demographic, biological, environmental and pregnancy-related predictors were included in the initial models. The complete list of assessed predictors can be found in Table S1 (Online Supplementary Document 1). Dummy variables indicating missingness for each predictor were included as additional variables, an approach that is justified for predictive models because it reflects the complete state of knowledge available at the time of prediction.

### Analysis

#### Descriptive statistics

A descriptive analysis of the background characteristics of women who experienced term compared to preterm delivery was performed using t-test for continuous variables, chi-square test for most of the binary variables and Fisher’s exact test for multiple gestations, to test for statistically significant differences between groups.

#### Prediction models

Five models were fit to predict risk of preterm delivery, including linear models and nonlinear decision tree approaches. All five strategies were designed to predict the outcome of preterm delivery using information available at 28 weeks of gestation. The first four models were time-to-event methods which modelled the time until delivery from the 28^th^ week gestation mark, accounting for left truncation and right censoring of person-time. Left truncation arises when women are enrolled beyond 28 weeks of gestation while right censoring arises when follow-up ceases prior to observation of the event of interest (e.g. due to outmigration or loss-to-follow-up). The first model was a Cox proportional hazards model, fit using the R package *survival* [21].

Second, an accelerated failure time model was fit with a log-logistic distribution using the R package *flexsurv* [22]. Third, a decision tree was fit using the R package *LTRtrees* that extends previous uses of a decision tree in survival analysis to account for left truncation and right censoring (LTRCART, left truncation right censoring classification and regression trees) [23]. Fourth, a decision tree ensemble was implemented using the *eXtreme Gradient Boosting* (XGB) R package which uses a Poisson likelihood function proposed by Fu and Simonoff (2017) to account for right censoring and left truncation [23,24]. Finally, a fifth analysis based on a XGB classification model was fit using a binary outcome (i.e. whether delivery was preterm or not) instead of the time-to-event. During fitting this last model, data which was either right censored or left truncated was excluded.

Models were fit and evaluated using 5-fold cross validation due to the need to evaluate models on out-of-sample data while reserving as much data as possible for fitting [25]. All models were evaluated on the same held-out dataset within each fold, regardless of which data or methods were used while fitting the model. Model performance was assessed using the AUC to assess accuracy at binary classification, and the c-index to assess the fraction of pairs for which predicted risk was concordant with delivery time. For both metrics, a value of 0.5 represents a random prediction which is uncorrelated with the true outcome. Larger values indicate more accurate predictions, and a value of 1 represents predictions which are perfectly concordant with the true outcome.

#### Simulation of cervical length and fetal fibronectin

A final analysis was performed to simulate the potential impact of including CL and FFN as predictors. These two variables were found in past work to be significantly associated with preterm birth [26-28]. Since they are not regularly collected in the study region, we used simulation to assess the potential gain from collecting them. The simulation used data from the MFMU PREDS study [29], a study which screened 2929 women for risk factors for preterm birth in the United States. PREDS study identified CL and FFN as key predictors for preterm birth [30,31]. Details on the simulation model and comparison between the simulated and real measurements can be found in the Supplemental Methods and Results (Online Supplementary Document 1).

## RESULTS

The sample composed 2834 pregnancies. Among those we excluded 75 (2.6%) records with gestational age at delivery <28 and ≥46 weeks, and a further 266 (9.4%) pregnancies whose follow-up did not go beyond 28^th^ gestational week. A total of 2493 pregnancies were included in the study. Of those, 138 (5.5%) women were lost to follow-up before delivery or did not have a recorded gestational age at delivery, so they were treated as censored observations in the time-to-event models and excluded from the binary classification model. A total of 968 (38.8%) women were enrolled in the cohort after 28 weeks of gestation (left-truncation), thus, time-varying predictors were considered missing for them since no information on those factors was available at the time of prediction. These women were also excluded from the binary classification model.

Among the 2355 women who were included in the study and followed until delivery, 14% had a preterm delivery. There was no difference in some background characteristics like age, body mass index, parity or history of previous preterm births among women with term deliveries compared to women with preterm deliveries (Table 1). However, the two groups showed significant differences in literacy (43.3% of women with term delivery were illiterate, compared to 50.5% of those who delivered prematurely), geographic location (42.8% of term deliveries occurred in the highland district within Birhan field site, compared to 52.4% of preterm deliveries), and multiple gestation (1.1% of term deliveries were multiple, compared to 3.4% of preterm deliveries).

**Table 1.** Characteristics of the study sample

| Variables | Total* |  |  | Term delivery |  |  | Preterm delivery |  |  |  |
| --- | --- | --- | --- | --- | --- | --- | --- | --- | --- | --- |
|  | N | mean | SD | N | mean | SD | N | mean | SD | p-value |
| Maternal age (years) | 2487 | 27.3 | 6.1 | 2030 | 27.3 | 6 | 319 | 27.2 | 6.4 | 0.69 |
| Body mass index (preconception) | 1108 | 22.0 | 3.1 | 930 | 22.0 | 3.2 | 138 | 21.9 | 2.6 | 0.60 |
|  | N | n | % | N | n | % | N | n | % | p-value |
| Illiterate | 2488 | 1105 | 44.4 | 2031 | 880 | 43.3 | 319 | 161 | 50.5 | 0.02 |
| Location: highland district (Angolela Tera) | 2489 | 1075 | 43.2 | 2032 | 870 | 42.8 | 319 | 167 | 52.4 | <0.01 |
| Primiparous | 2493 | 793 | 31.8 | 2034 | 627 | 30.8 | 321 | 108 | 33.6 | 0.34 |
| History of a previous preterm birth | 2493 | 47 | 1.9 | 2034 | 38 | 1.9 | 321 | 7 | 2.2 | 0.87 |
| Multiple gestation | 2408 | 35 | 1.5 | 2034 | 23 | 1.1 | 321 | 11 | 3.4 | <0.01 |
| Neonatal sex: female | 2355 | 1144 | 48.6 | 2034 | 993 | 48.8 | 321 | 151 | 47.0 | 0.59 |

\*Total counts include censored study participants with unknown pregnancy outcome or date of delivery
SD – Standard Deviation

Some predictors had high levels of missing data, particularly where our study relied on facility visits. Approximately 25% of participants did not attend any antenatal care visit in the study health facilities after being enrolled in the cohort, thus, variables collected at antenatal care visits, such as current infections or concomitant diseases were missing for these women. Further, over 70% of women who attended at least one antenatal care visit had missing data on lab and point-of-care results such as white blood cell counts, proteinuria or bacteriuria.

The predictive performance of all models was generally poor (Table 2). The c-statistic and AUC were highest for the XGB classification model, with an AUC of 0.60. The receiver operating characteristic curves (ROC) depict the trade-off between the false and true positive rates achieved by varying the threshold for classifying delivery as preterm or term (Figure 1). As an example, at the point on this curve corresponding to a 90% true positive rate, all models had a false positive rate of at least 75%, indicating a lack of specificity in picking out women who are truly at higher risk.

**Table 2.** Performance metrics of the different predictive models

| Model | AUC (95%CI) |  | c statistic (95% CI) |  |
| --- | --- | --- | --- | --- |
| Accelerated failure time | 0.57 | (0.54, 0.61) | 0.53 | (0.52, 0.55) |
| Cox | 0.51 | (0.47, 0.54) | 0.52 | (0.50, 0.53) |
| LTRCART | 0.54 | (0.51, 0.58) | 0.53 | (0.51, 0.55) |
| XGBoost (classification) | 0.60 | (0.57, 0.63) | 0.54 | (0.52, 0.55) |
| XGBoost (survival) | 0.52 | (0.49, 0.56) | 0.52 | (0.50, 0.53) |

AUC – Area Under the receiver operating characteristic Curve; CI – Confidence Interval; LTRCART – Left Truncation Right Censoring Classification And Regression Trees; XGBoost – eXtreme Gradient Boosting

**Figure 1.**
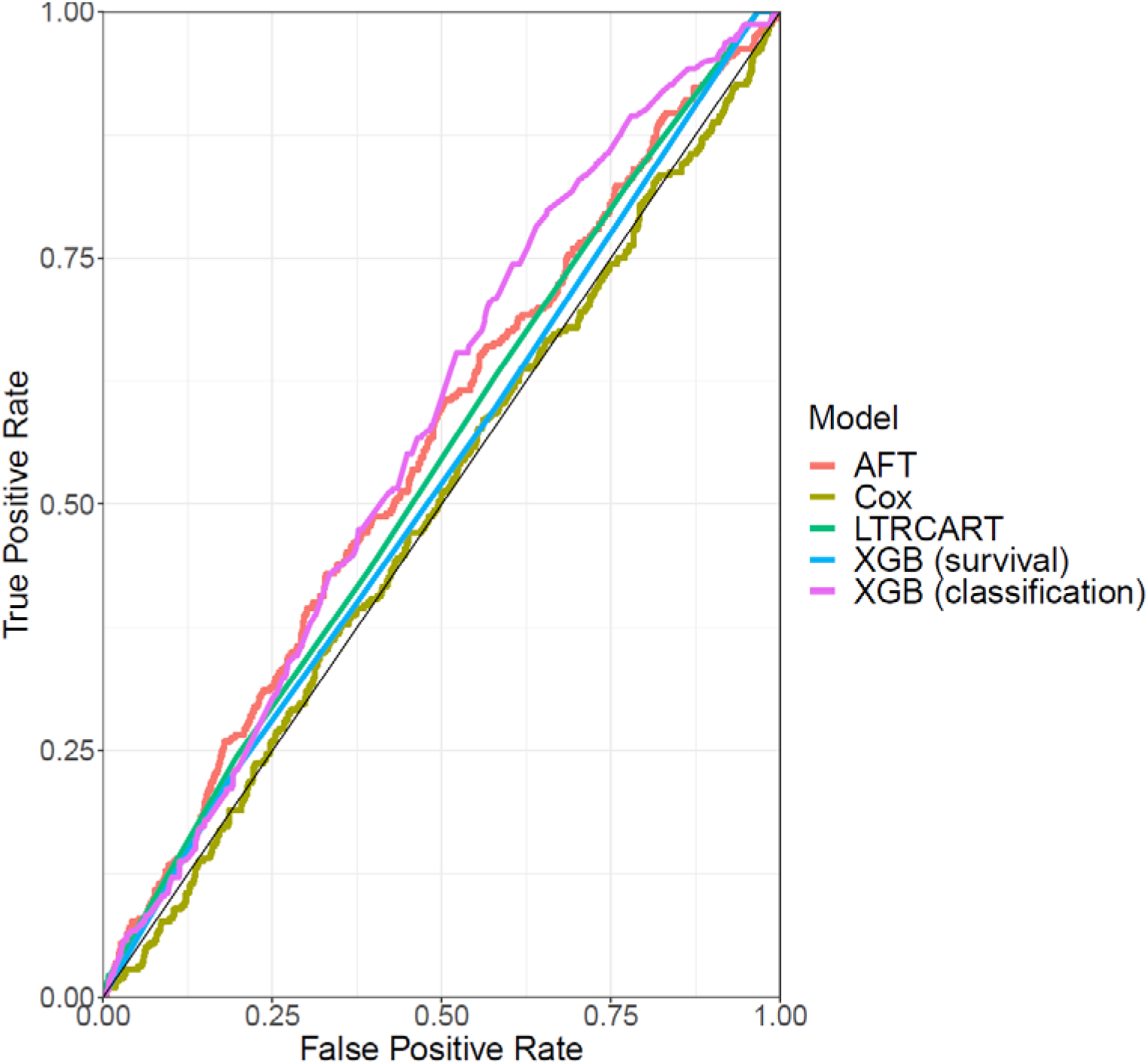
ROC curves for each model. AFT – Accelerated Failure Time; LTRCART – Left Truncation Right Censoring Classification And Regression Trees; ROC - Receiver Operating Characteristic Curve; XGB – eXtreme Gradient Boosting

There was substantial heterogeneity in the factors that were ultimately retained in the five models (Table 3). Both biological and socio-demographic factors were among the top contributors of standard time-to-event models. Regarding decision tree models, the top five predictors are mainly biological, with neonatal sex being the predictor with the greatest importance.

**Table 3.** Top 5 predictors for each model: model coefficients and feature importance scores

| Model | Top predictors |  |  |  |  |
| --- | --- | --- | --- | --- | --- |
|  | 1 | 2 | 3 | 4 | 5 |
| Accelerated failure time:<br>*Accelerator factor* | Multiple gestation<br>0.74 | Lymphocyte % = NA<br>1.26 | Vaginal bleeding<br>1.22 | History of pre-eclampsia<br>0.83 | Maternal age = NA<br>0.83 |
| Cox:<br>*Hazard ratio (95%CI)* | Multiple gestation<br>3.15 (0.61, 16.21) | History of diabetes<br>0.33 (0.03, 3.61) | Maternal age = NA<br>2.79 (1.72, 4.55) | Lymphocyte % = NA<br>2.68 (0.10, 1.63) | History of pre-eclampsia<br>2.29 (1.44, 3.62) |
| LTRCART:<br>*Split level* | Neonatal sex<br>1 | Multiple gestation<br>2 | NA<br>- | NA<br>- | NA<br>- |
| XGBoost classification:<br>*Gain* | Location (district)<br>0.11 | Parity<br>0.10 | Family size = top quintile<br>0.10 | Neonatal sex<br>0.09 | Systolic blood pressure<br>0.08 |
| XGBoost survival:<br>*Gain* | Neonatal sex<br>1.00 | NA<br>- | NA<br>- | NA<br>- | NA<br>- |

*LTRCART* – Left Truncation Right Censoring Classification And Regression Trees; *NA* – Not Applicable/missing data; *XGBoost* – eXtreme Gradient Boosting

The performance of several individual models improved when simulated measurements of CL and FFN were included as features for each individual, particularly the accelerated failure time and LTRCART decision tree models (Table 4). However, no model exceeded an estimated AUC of 0.60 indicating that the overall predictability of preterm delivery did not change substantially from the inclusion of these additional predictors.

**Table 4.** Performance metrics of the models with simulated measurements

| Model | AUC (95%CI) |  | c statistic (95% CI) |  |
| --- | --- | --- | --- | --- |
| Accelerated failure time | 0.60 | (0.56, 0.63) | 0.55 | (0.53, 0.57) |
| Cox | 0.55 | (0.52, 0.59) | 0.53 | (0.50, 0.55) |
| LTRCART | 0.60 | (0.56, 0.64) | 0.55 | (0.52, 0.57) |
| XGBoost (classification) | 0.60 | (0.56, 0.64) | 0.55 | (0.52, 0.56) |
| XGBoost (survival) | 0.48 | (0.45, 0.51) | 0.50 | (0.48, 0.51) |

*AUC* – Area Under the receiver operating characteristic Curve; *CI* – Confidence Interval; *LTRCART* – Left Truncation Right Censoring Classification And Regression Trees; *XGBoost* – eXtreme Gradient Boosting

## DISCUSSION

Our study shows that risk prediction of preterm delivery remains a challenge in the absence of data on biomarkers. Despite using a wide range of methodological approaches to adjust for missing data, competing risk of stillbirths, and late cohort enrollments, both traditional epidemiological and machine learning models performed poorly and had low specificity in identifying women who delivered before 37 weeks of gestation. This low predictive performance differs with existing models with higher predictive ability that were designed to predict preterm birth among women at an already high risk due to obstetric conditions such as twin pregnancy [7,8], short cervix, cervical insufficiency [10,32], or hospital admission due to preterm labor [6,33,34]. Other higher performing algorithms used predictors that were not available or applicable in low-resource settings such as amniotic and cervical fluids [33], inflammatory markers [35], or method of conception [5,8]. Lack of a fixed prediction time point and competing risk of stillbirth are methodological gaps of most published studies.

To our knowledge, our study is among the few in developing risk prediction models in a low-resource setting. Only one published study presented the development of a model for preterm birth prediction in Ethiopia [36]. Despite reporting good model performance, this study presents an important limitation; it predicts preterm birth retrospectively using all information available in hindsight (e.g. events such as premature rupture of membranes), while our aim is to assess whether early prediction is possible to inform preventive interventions, leading us to fix a time point for prediction (28 weeks of gestation). Moreover, their study was conducted using data from a hospital-based cohort, likely to be composed of women at higher *a priori* risk of adverse outcomes.

Studies carried out in Ethiopia identified risk factors for preterm delivery including obstetric conditions, socio-demographic characteristics, urinary and vaginal infections, and hypertensive disorders [37,38]. We included all these factors to build the most accurate models with the available data. Among all predictors, neonatal sex was assigned high importance in the decision tree models despite the small difference in prevalence between boys and girls. This is consistent with higher rates of preterm birth for male fetuses in other studies [39,40]. Nevertheless, prediction studies like this aim to characterize prognosis and to anticipate or forecast an outcome.

We simulated the conditional distribution of CL and FFN to test whether access to these measurements may improve models’ predictive ability. However, the improvement of the models was negligible, indicating that women who are identified as high-risk via CL or FFN could also be identified as high risk via other predictors. Although both CL and FFN are among the most used indicators to identify high risk pregnancies for preterm delivery in clinical practice, their measurement is not always recommended in *a priori* low-risk populations [15,41]. Our results do not support the allocation of resources to CL and FFN measurement in low-resource settings to predict risk of preterm birth. Similarly, other studies observed a poor predictive power of CL and FFN in the absence of additional maternal predictors [42].

The results of this study have important research-related and public health implications. Given the poor performance of all available predictive models, it is fundamental to continue research on the underlying causes of preterm delivery, better understanding the pathways between different risk factors and preterm birth, in order to predict and prevent preterm births in the future. Most cases still occur among women without any known risk factor [14,15]. It is crucial to look for new indicators and biomarkers of preterm delivery. Genetic factors, immunological biomarkers and protein expression are showing promising results [43,44]. There may be value in exploring the use of ‘omics’ since no biomarkers predictive of preterm birth have yet been identified [45]. While all settings can benefit from such technologies, from an equity perspective it may be especially important to ensure availability in low-resource settings where the survival of preterm infants is lower and identifying high-risk women can enable targeted preventative interventions.

Predictive algorithms with modest performance could be used to identify pregnancies at a very low risk of preterm delivery, thus excluding them from interventions. However, a large proportion of women at a low risk will still be targeted in those interventions due to the low specificity of the models. In Ethiopia, recommending pregnant women to stay in maternity waiting homes is part of the birth preparedness strategy, though it has not been demonstrated to improve pregnancy outcomes [46,47]. Targeting the recommendation of staying in maternity waiting homes to a reduced number of individuals would increase the cost-effectiveness of the intervention and improve the pregnancy experience of some women.

Our findings should be interpreted considering some limitations. Similar to most longitudinal studies, there was study attrition. To address loss to follow-up, we adjusted time-to-event models for censoring and created a “missing” category for all predictors with missing data. The use of a composite outcome that included all preterm deliveries regardless of vital status of the fetus or neonate did not enable the models to separately predict the risk of having a live preterm baby from the risk of having a preterm stillbirth. However, the use of a combined outcome allows us to address competing risks of preterm stillbirths, a common limitation of other available prediction models. Despite these limitations, our study fills an evidence gap by exploring prediction of preterm delivery during the first 28 weeks of gestation in a resource-limited setting with important restrictions in data availability. We considered a comprehensive selection of >70 predictors and tested five different algorithms: Accelerated Failure Time, Cox, LTRCART and two decision tree ensembles. We acknowledge that the classification tree ensemble is not recommended for data with censoring or truncation. However, we fit this model together with the other four algorithms for the purpose of being fully exhaustive in our effort to explore all potential methodological options to achieve our study aim of developing an accurate algorithm. The results of all of them show that the difficulty of predicting preterm delivery is robust to potential variation in the process of constructing risk models.

In settings with low coverage of antenatal care and limited resources to perform ultrasound and biomarker measurements, predicting risk of preterm delivery remains a major challenge. New indicators of preterm delivery may be necessary to enable targeted interventions.

## Supporting information

TRIPOD Checklist

## Data Availability

Data are available upon reasonable request. Data use is governed by the Birhan Data Access Committee (DAC) and follows Birhan's data sharing policy. All researchers who wish to access Birhan data can complete a Birhan data request form and submit it for decision by the Birhan DAC. Datasets will only be provided with deidentified data to maintain confidentiality of study participants.

## AUTHORSHIP CONTRIBUTIONS

CPD, BW, SH and GJC conceptualized and designed the study. GJC is the study PI, she obtained funding for the study, and supervised all study activities. DB is the co-PI of the study, he helped to obtain funding for the study and led data collection team. BMH, DB and GJC participated in data collection. CPD and BW conducted the data analysis. SH oversaw the data analysis. FGBG curated the data for this study. CPD and BW drafted the first version of the manuscript. All authors critically revised the manuscript for important intellectual content. All authors approved the final version of the manuscript.

## ACKNOWLEDGMENTS

We thank all the mothers and children who participated in the study (HDSS and MCH cohort) and the community of the Birhan field site. We also thank data collectors, supervisors, coordinators, and the HaSET team for their contributions.

## ETHICAL CONSIDERATIONS

Ethical clearance was obtained from the Ethics Review Board (IRB) of Saint Paul’s Hospital Millennium Medical college, (Addis Ababa, Ethiopia), and Harvard T.H. Chan School of Public Health (Boston, United States). Signed informed consent was obtained from all participants.

## SUPPLEMENTARY DOCUMENTS

### Online Supplementary Document 1

Table S1. Predictors assessed for inclusion in the models

Supplemental Methods and Results. Simulation of cervical length and foetal fibronectin: methods and fit between simulated and real values:

Table S2. Coefficients for simulated model of cervical length and foetal fibronectin

Figure S1. Joint distribution of gestational age and cervical length in the PREDS dataset (left) and simulated values (right). The blue line gives a LOWESS smoothing

Figure S2. Joint distribution of foetal fibronectin (1: positive, 0: negative) and cervical length in the PREDS dataset (left) and simulated values (right). The blue line gives a LOWESS smoothing

**TRIPOD Checklist**

