## Supplementary material for "Development of risk prediction models for preterm delivery in a rural setting in Ethiopia": Online Supplementary Document 1

**Table of contents**

**Table S1**. Predictors assessed for inclusion in the models, pages 2-11

**Supplemental Methods and Results**. Simulation of cervical length and fetal fibronectin: methods and fit between simulated and real values, pages 12-13

**Table S2.** Coefficients for simulated model of cervical length and fetal fibronectin, page 12

**Figure S1.** Joint distribution of gestational age and cervical length in the PREDS dataset (left) and simulated values (right). The blue line gives a LOWESS smoothing, page 13

**Figure S2.** Joint distribution of fetal fibronectin (1: positive, 0: negative) and cervical length in the PREDS dataset (left) and simulated values (right). The blue line gives a LOWESS smoothing, page 13

**Table S1. Predictors assessed for inclusion in the models**

| **Predictors** | **Included in the models** | **Reasons for exclusion** | **Other remarks** |
| --- | --- | --- | --- |
| **Socioeconomic and demographic** | | | |
| Maternal age | Yes |  |  |
| Maternal education | Yes |  |  |
| Maternal literacy | Yes |  |  |
| Marital status | Yes |  |  |
| Family has own income | Yes |  |  |
| Main source of income | Yes |  |  |
| Wealth index | Yes |  |  |
| Family size (number of individuals in the household) | Yes |  |  |
| Ethnicity | Yes |  |  |
| Religion | Yes |  |  |
| Urban/rural area | Yes |  | Woreda used as a proxy |
| Altitude | Yes |  | Woreda used as a proxy |
| Time and distance to the nearest health facility | Yes |  |  |
| **Environmental and behavioral** | | | |
| Household air pollution | No | Not available in the study database |  |
| Alcohol intake | Yes |  |  |
| Tobacco use | No | No smokers in the sample |  |

**Table S1. Predictors assessed for inclusion in the models (continued)**

| Substance abuse | | No | Rare event (≤5 cases) |  |
| --- | --- | --- | --- | --- |
| Iron intake | | Yes |  |  |
| Folic acid intake | | Yes |  |  |
| Mebendazole intake | | Yes |  |  |
| Other medication intake | | Yes |  |  |
| Multivitamin use | | Yes |  |  |
| Vitamin A intake | | Yes |  | Intake of Vitamin A rich fruits and vegetables |
| Zinc intake | | No | Not available in the study database |  |
| Oral rehydration salts | | No | Not available in the study database |  |
| Nutrition: intake of different products (fruits, vegetables, oil, coffee, meet, eggs, fish) | | Yes |  |  |
| Fasting during pregnancy | | Yes |  |  |
| Insecticide-treated nets use | | Yes |  |  |
| **Anthropometric measurements** | | | | |
| Height | Yes | |  | Preconception measurement |
| Weight | Yes | |  | Preconception measurement |
| Body Mass Index | Yes | |  | Preconception measurement |
| Mid upper arm circumference | Yes | |  | Measurement from enrolment visit used as a proxy of preconception measurement |

**Table S1. Predictors assessed for inclusion in the models (continued)**

| **History of previous diseases and conditions** | | | |
| --- | --- | --- | --- |
| Diabetes | Yes |  |  |
| Eclampsia | Yes |  | Combined with pre-eclampsia |
| Pre-eclampsia | Yes |  | Combined with eclampsia |
| Sexually transmitted diseases | Yes |  |  |
| Cardiac disease | No | Rare event (≤5 cases) |  |
| Chronic hypertension | No | Rare event (≤5 cases) |  |
| Renal disease | No | Rare event (≤5 cases) |  |
| Severe medical disease or condition | No | Rare event (≤5 cases) |  |
| **Obstetric history** | | | |
| Previous multiple gestation | Yes |  |  |
| Previous stillbirth | Yes |  |  |
| Previous miscarriage | Yes |  |  |
| Previous preterm delivery | Yes |  |  |
| Previous low birth weight neonate | Yes |  |  |
| Previous newborn with birth defect | No | Rare event (≤5 cases) |  |
| Previous caesarean section | Yes |  |  |
| Previous antepartum hemorrhage | No | Not available in the study database |  |
| Interpregnancy interval | Yes |  |  |

**Table S1. Predictors assessed for inclusion in the models (continued)**

| Parity | Yes |  |
| --- | --- | --- |
| Gravidity | Yes |  |
| History of surgeries in reproductive tract | Yes |  |
| Method of conception | No | Not available in the study database |
| **Current infections and concomitant diseases** | | |
| Amniotic fluid infection | No | Not available in the study database |
| Bacterial vaginosis | No | Not available in the study database |
| Chlamydia | No | Not available in the study database |
| Maternal chorioamnionitis | No | Rare event (≤5 cases) |
| Syphilis | Yes |  |
| Pelvic infection | No | Not available in the study database |
| Malaria | No | Not available in the study database |
| HIV | Yes |  |
| Hepatitis B | No | Rare event (≤5 cases) |
| Sickle cell anemia | No | Not available in the study database |
| Eclampsia | No | Rare event (≤5 cases) |

**Table S1. Predictors assessed for inclusion in the models (continued)**

| Pre-eclampsia | No | No cases of pre-eclampsia in the sample | |
| --- | --- | --- | --- |
| Asthma | No | Not available in the study database | |
| Diabetes | No | Only 2 women screened for diabetes | |
| Mental disorder | No | Not available at origin (week 28 of gestation) | |
| Thyroid disease | No | Not available in the study database | |
| Pregnancy induced hypertension | No | Rare event (≤5 cases) | |
| Psychological and social stress | No | Not available in the study database | |
| Chest abnormality | No | Rare event (≤5 cases) | |
| Heart abnormality | No | Rare event (≤5 cases) | |
| Depression | No | Not available at origin (week 28 of gestation) | |
| **Antenatal care** | | | |
| Number of antenatal care visits | Yes |  |  |
| Early antenatal care attendance | Yes |  | Two predictors considered: first ANC visit before week 13 and before week 24 |

**Table S1. Predictors assessed for inclusion in the models (continued)**

| **Biological, signs and symptoms** | | | |
| --- | --- | --- | --- |
| Multiple gestation | Yes |  | Information collected at delivery. Suspected multiple gestation for women who were lost to follow-up |
| Neonatal sex | Yes |  | Information collected at delivery. Unknown for women who were lost to follow-up |
| Preterm labor | No | Rare event (≤5 cases) |  |
| Premature rupture of membranes | No | Rare event (≤5 cases) |  |
| Vaginal bleeding | Yes |  |  |
| Antepartum hemorrhage | No | Rare event (≤5 cases) |  |
| Vaginal discharge syndrome | No | Rare event (≤5 cases) |  |
| Genital ulcer | No | Rare event (≤5 cases) |  |
| Pallor | Yes |  |  |
| Jaundice | Yes |  |  |
| Severe nausea and vomiting | Yes |  |  |
| Headache | Yes |  |  |
| Blurred vision | Yes |  |  |
| Right upper quadrant abdominal pain | Yes |  |  |
| Fever | Yes |  |  |
| Pain during urination | Yes |  |  |

**Table S1. Predictors assessed for inclusion in the models (continued)**

| Increased urinary frequency | Yes |  |  |
| --- | --- | --- | --- |
| Increased urinary urgency | Yes |  |  |
| Constipation | No | Rare event (≤5 cases) |  |
| Painful sexual intercourse | No | Not available in the study database |  |
| Seizure | No | Not available in the study database |  |
| Fetal movement recall | Yes |  |  |
| Composite predictor: Danger signs and symptoms | Yes |  | At least one of the following signs and symptoms: headache, blurring of vision, vaginal bleeding, urinary pain, urinary frequency, urinary urgency, decreased foetal movement, vaginal discharge syndrome, right upper quadrant abdominal pain, fever, pallor, jaundice, severe vomiting or nausea |
| Composite predictor: Severe maternal complications | No | Rare event (≤5 cases) | At least one of the following maternal complications: premature rupture of membranes, antepartum haemorrhage and pregnancy induced hypertension |
| **Ultrasound measurements** | | | |
| Cervical length (CL) | No | Not available in the study database |  |

**Table S1. Predictors assessed for inclusion in the models (continued)**

| Cervical glandular area | No | Not available in the study database |
| --- | --- | --- |
| Foetal breathing movements | No | Not available in the study database |
| Biometry | No | Not available in the study database |
| Intrauterine growth restriction | No | Not available in the study database |
| **Other laboratory, non-laboratory, and point-of-care measurements** | | |
| Foetal heartbeat | No | Not available at origin (week 28 of gestation) |
| Foetal respiratory rate | No | Not available in the study database |
| 0_2_ saturation | No | Not available in the study database |
| Blood pressure | Yes |  |
| Hb / anaemia | Yes |  |
| Haematocrit | Yes |  |
| Blood glucose levels | No | Only 2 women with measured blood glucose levels |
| Blood group | Yes |  |
| Rh factor | Yes |  |
| White blood cells (in x1000 cells/µl) | Yes |  |
| Neutrophils (%) | Yes |  |

**Table S1. Predictors assessed for inclusion in the models (continued)**

| Lymphocytes (%) | Yes |  |
| --- | --- | --- |
| Nitrites in urine | No | Rare event (≤5 cases) |
| Leukocyte esterase in urine | Yes |  |
| Bacteriuria | Yes |  |
| Proteinuria | Yes |  |
| Micronutrient levels in blood | No | Not available in the study database |
| Hormone levels (cortisol, corticotropin-releasing hormone, salivary estriol, human chorionic gonadotropin, relaxin) | No | Not available in the study database |
| Fetal Fibronectin (FFN) | No | Not available in the study database |
| Alpha fetoprotein level | No | Not available in the study database |
| Pregnancy-associated plasma protein A (PAPP-A) | No | Not available in the study database |
| Interleukin levels | No | Not available in the study database |
| Vaginal microbiota | No | Not available in the study database |
| Other cervical, amniotic fluid and blood biomarkers (alkaline phosphatase, 𝛽-2-microglobulin, C-reactive protein, defensins, IGFBP-1, matrix metalloproteinase-8, granulocyte colony stimulating factor) | No | Not available in the study database |

**Table S1. Predictors assessed for inclusion in the models (continued)**

| Genetic polymorphisms | No | Not available in the study database |
| --- | --- | --- |

**Note:** all predictors included in the models were generated with information available at origin/time of prediction (week 28 of gestation), or using data collected later in pregnancy only if the predictor was time-invariant.

**Supplemental Methods and Results. Simulation of cervical length and fetal fibronectin: methods and fit between simulated and real values**

The simulation modelled the joint distribution of cervical length (CL) and fetal fibronectin (FFN) conditioned on the true gestational age of the delivery, to capture the fact that both variables contain (potentially noisy) information about the outcome of interest. We modelled CL as a continuous variable and FFN as a binary variable (i.e., a positive or negative test result). Their joint distribution was also conditioned on age, parity, and body mass index (BMI), three variables associated with CL and FFN which are present in both datasets. This helped adjust the simulation model to account for differences between the two populations. A factorization of the joint distribution was specified as

$$\Pr\left[ CL, FFN \right]=\Pr\left[ CL \right]\cdot\Pr\left[ FFN | CL \right]=N\left( \beta_{CL,\mu^{\chi}}, \beta_{CL,\sigma^{\chi}} \right)\cdot\sigma(\beta_{FFN}\left[ \chi CL \right])$$

where $N$ is the normal distribution, σ is the inverse logit function and the β are parameters which control the mean and variance of each term. The model was fit using maximum likelihood on the PREDS dataset. Table S2 shows the fitted values for each coefficient. Measurements for CL and FFN were simulated from the model for our MCH cohort using these inferred parameters. To assess the fidelity of the simulation, we also simulated measurements for the PREDS cohort and compared the joint distribution of gestational age and each measurement in the PREDS dataset versus simulation. Figure S1 shows the real and simulated joint distribution of CL and gestational age. Figure S2 shows the real and simulated joint distribution of FFN and gestational age. We observe a high degree of consistency between the real and simulated joint distributions for both measurements, though some discrepancies are present in the conditional mean of the right tail of the FFN-gestational age distribution. This is due to the very small number of data points which fall into this part of the distribution.

**Table S2. Coefficients for simulated model of cervical length and foetal fibronectin.**

| **Variable** | $\boldsymbol{\beta}_{\boldsymbol{CL, \mu}}$ | $\boldsymbol{\beta}_{\boldsymbol{CL, \sigma}}$ | $\boldsymbol{\beta}_{\boldsymbol{FFN}}$ |
| --- | --- | --- | --- |
| Gestational age (weeks) | 0.629 | -0.247 | -0.211 |
| Age (years) | 0.057 | 0.051 | 0.007 |
| Parity | 2.000 | 0.315 | 0.306 |
| BMI | 0.088 | 0.039 | 0.020 |
| Cervical length (mm) | N/A | N/A | -0.058 |
| Intercept | 6.036 | 15.087 | 5.461 |

BMI – Body Mass Index; CL – Cervical Length; FFN – Foetal Fibronectin

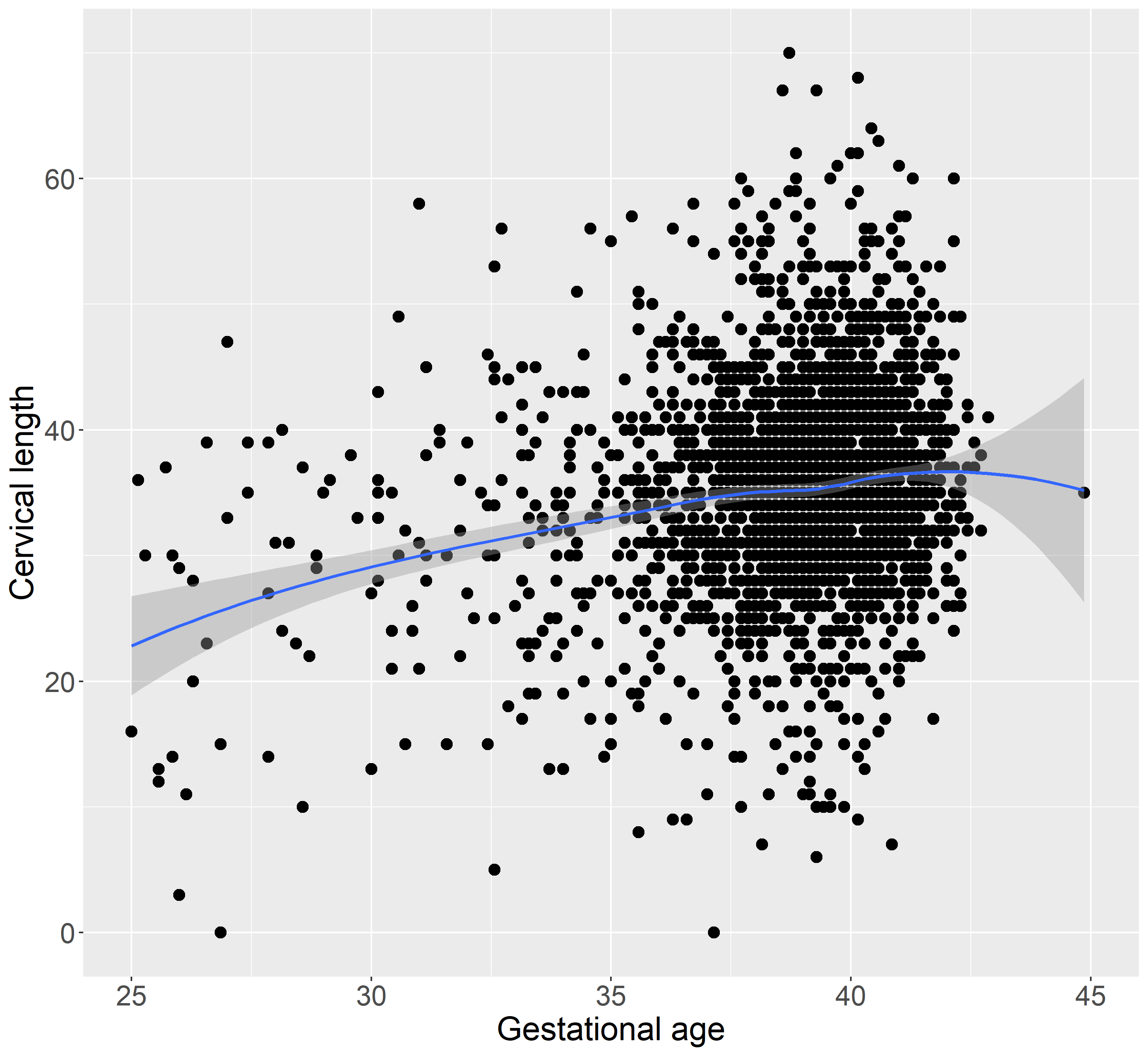

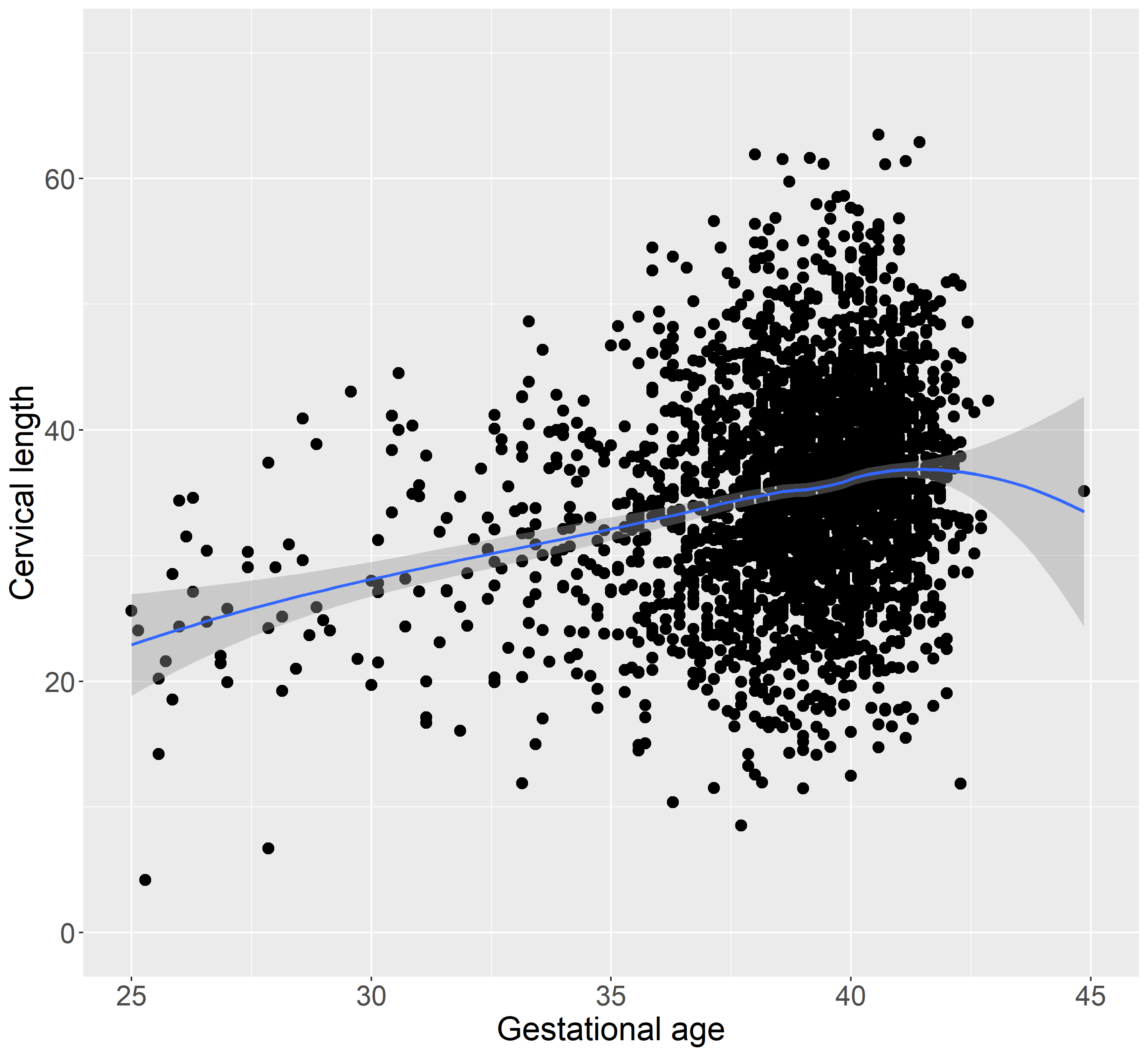

**Figure S1. Joint distribution of gestational age and cervical length in the PREDS dataset (left) and simulated values (right). The blue line gives a LOWESS smoothing.**

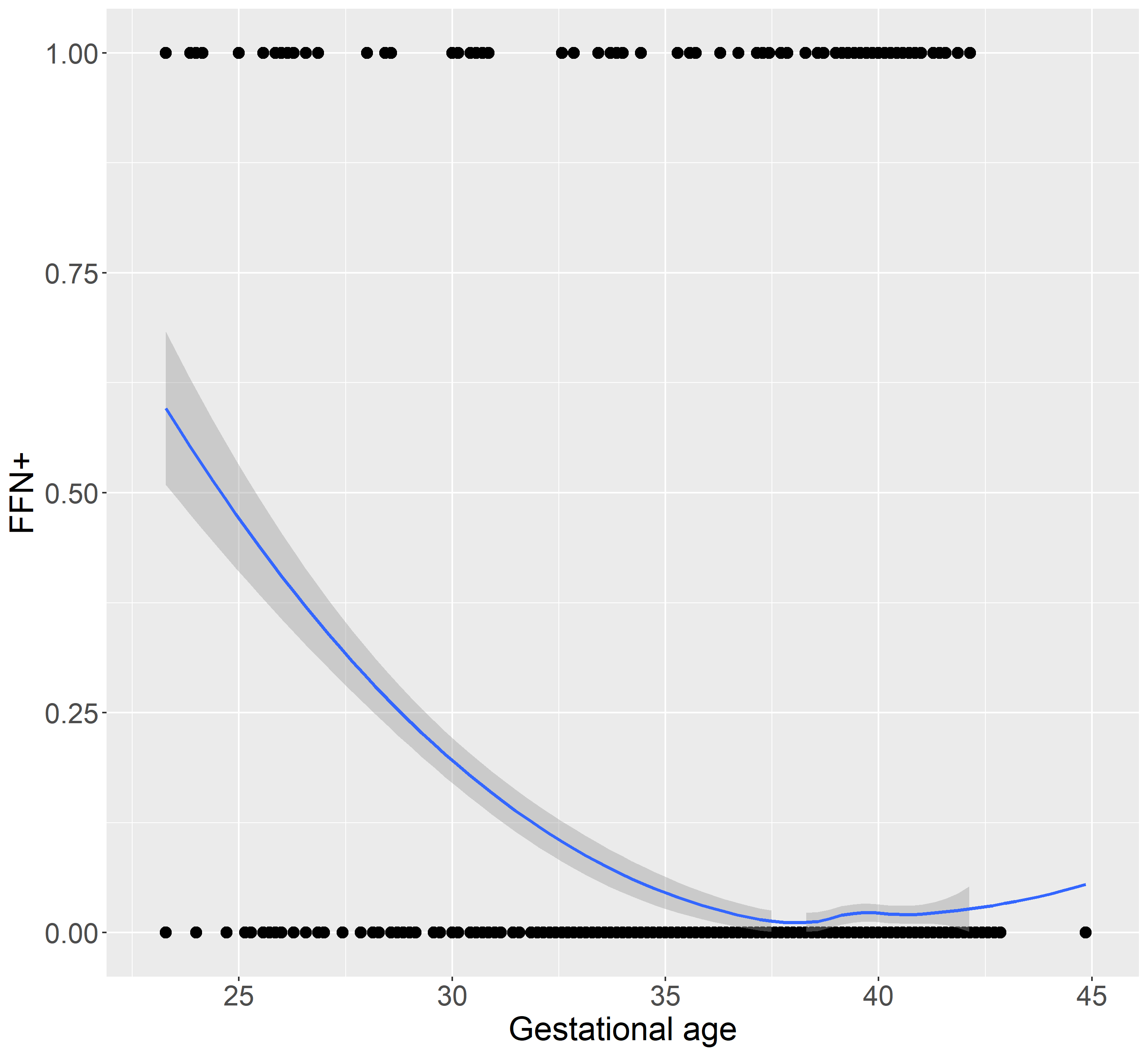

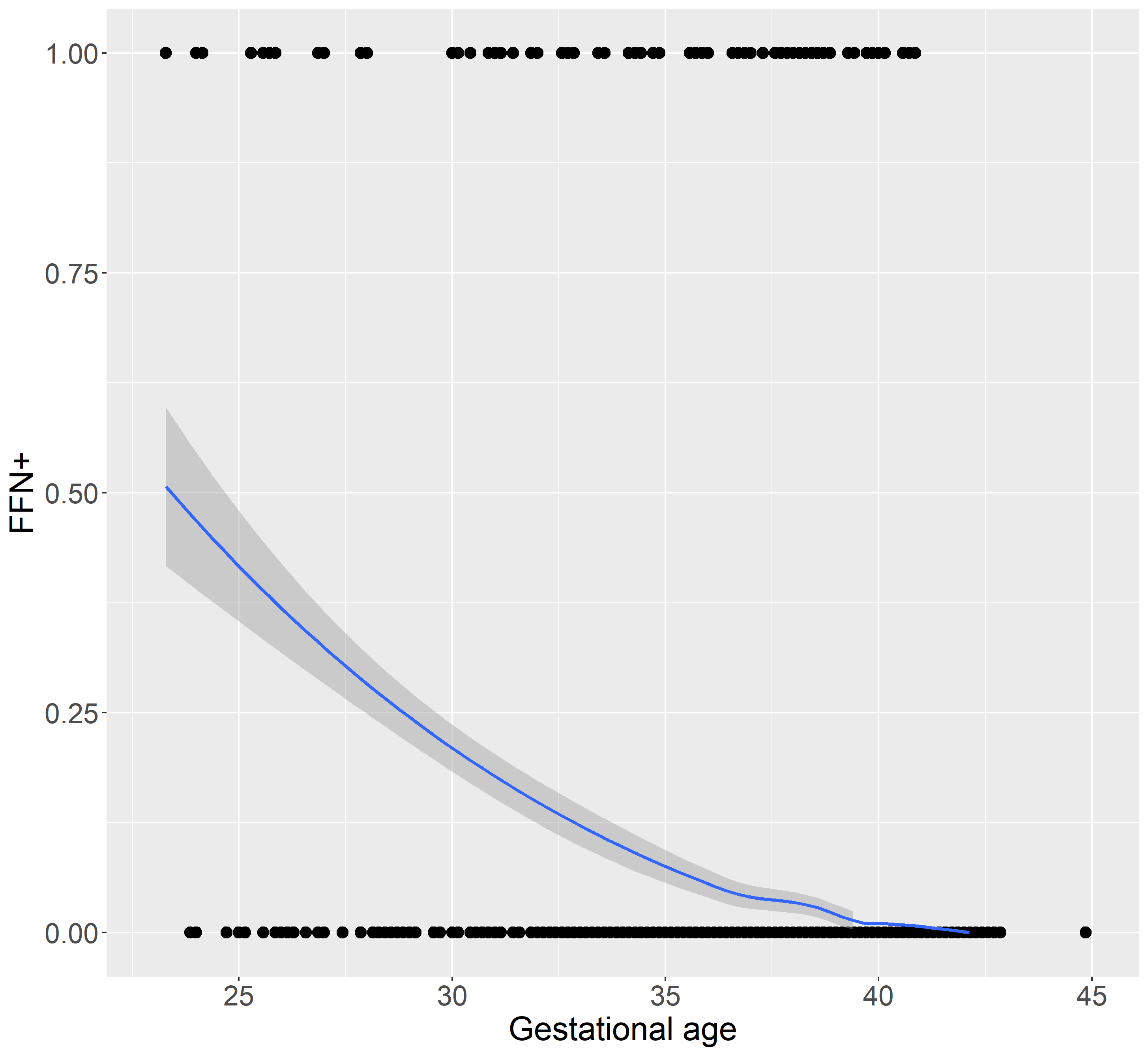

**Figure S2. Joint distribution of fetal fibronectin (1: positive, 0: negative) and cervical length in the PREDS dataset (left) and simulated values (right). The blue line gives a LOWESS smoothing.**
